# Bayesian hierarchical modeling of joint spatiotemporal risk patterns for Human Immunodeficiency Virus (HIV) and Tuberculosis (TB) in Kenya

**DOI:** 10.1101/2020.01.22.20018390

**Authors:** Verrah A. Otiende, Thomas N. Achia, Henry G. Mwambi

## Abstract

The spatiotemporal modeling of multiple diseases simultaneously is a recent extension that advances the space-time analysis to model multiple related diseases simultaneously. This approach strengthens inferences by borrowing information between related diseases. Numerous research contributions to spatiotemporal modeling approaches exhibit their strengths differently with increasing complexity. However, contributions that combine spatiotemporal approaches to modeling of multiple diseases simultaneously are not so common. We present a full Bayesian hierarchical spatio-temporal approach to the joint modeling of Human Immunodeficiency Virus and Tuberculosis incidences in Kenya. Using case notification data for the period 2012 - 2017, we estimated the model parameters and determined the joint spatial patterns and temporal variations. Our model included specific and shared spatial and temporal effects. The specific random effects allowed for departures from the shared patterns for the different diseases. The space-time interaction term characterized the underlying spatial patterns with every temporal fluctuation. We assumed the shared random effects to be the structured effects and the disease-specific random effects to be unstructured effects. We detected the spatial congruence in the distribution of Tuberculosis and Human Immunodeficiency Virus in approximately 29 counties around the western, central and southern regions of Kenya. The distribution of the shared relative risks had minimal difference with the Human Immunodeficiency Virus disease-specific relative risk whereas that of Tuberculosis presented many more counties as high-risk areas. The flexibility and informative outputs of Bayesian Hierarchical Models enabled us to identify the similarities and differences in the distribution of the relative risks associated with each disease. Estimating the Human Immunodeficiency Virus and Tuberculosis shared relative risks provide additional insights towards collaborative monitoring of the diseases and control efforts.

## Introduction

Disease mapping as a modeling approach is commonly used to describe the geographical distribution of disease burden thereby generating hypotheses on their possible causes and differences [1]. Statistical methods for characterization of the geographical variations have contributed significantly towards advancing the focus of disease mapping to incorporate other analysis techniques. Including the time dimension has given rise to the spatiotemporal modeling of the variation of disease risk which enables simultaneously studying the spatial patterns and temporal variations thereby giving deeper insights over purely spatial mapping [2,3]. Another extension is the joint spatial modeling of multiple related diseases with common risk factors, which outspread the standard univariate disease mapping methodologies. It generates both the shared and divergent trends thereby increasing the estimation precision of disease risk [4]. The latest and most recent extension is the joint spatiotemporal modeling of multiple diseases. This approach advances the space-time analysis to model multiple diseases simultaneously thereby strengthening inference by borrowing information across neighboring regions and between related diseases or sub-populations with common aetiological factors [5]. The benefits of borrowing information lie in the ability to observe concurrency of patterns and to allow conditioning of one disease on others [6] which is very valuable when accounting for uncertainty due to sparse disease count or underreporting [7]

Studies combining spatiotemporal approaches to modeling multiple diseases simultaneously are not so common despite the development and application of novel computational techniques. Reviews from literature show few contributions of these approaches that exhibit their strengths differently with increasing complexities. [8] used a Bayesian factor analysis approach to combine the space-time disease mapping and joint modeling of different cancers. [9] used a full Bayesian hierarchical model to split the disease risks into shared and disease-specific spatiotemporal components. Their definition of multiple diseases was male and female subpopulations. [10] also applied a full Bayesian hierarchical approach to their spatiotemporal model to estimate the relative risk of various cancers while adjusting for age and gender. Using the hierarchical Bayesian factor models, [11] combined the dynamic factor analytic models with space-time disease mapping and produced a flexible framework for jointly analyzing multiple related diseases. Another study by [12] compared three formulations of the spatiotemporal shared component model on five diseases and examined the changes in the shared factors over time.

Incorporating the spatiotemporal modeling approaches to modeling of multiple diseases simultaneously adds significant complexity to the model structure [13]. The Bayesian hierarchical modeling approach makes an appropriate framework for solving complexities in the spatiotemporal structures [9,10,14,15]. The uniqueness of the Bayesian approach that differentiates it from other classical approaches is its robustness to interpretation of the posterior estimates and generating inferences of all model parameters [16].

The feasibility of using case notifications as a surrogate of population-based studies to estimate the shared and disease-specific risks of multiple diseases is also unknown. Against this background, we investigate the spatial and temporal patterns of Human Immunodeficiency Virus (HIV) and Tuberculosis (TB) burden in Kenya jointly using case notification data for a six-year period and characterize the areas with unusually high relative risks. These model maps describe new exposure hypotheses that warrant further epidemiological investigations on existing challenges and opportunities for disease surveillance and etiology

### Motivation of the study

The HIV and TB diseases have a co-epidemic overlap in their epidemiologic characteristics and clinical manifestations [17]. Both the HIV and TB pathogens interact synergistically, accelerating the progress of illness thereby increasing the likelihood of death [18]. TB is a communicable disease and a major cause of ill health globally that affects the lungs (pulmonary) but can also affect other sites (extrapulmonary) [19,20]. Despite being a curable and preventable disease, TB is the leading cause of death from bacterial infection worldwide and undoubtedly representing a global public health priority [21–24]. TB is also the leading cause of morbidity and mortality among people living with HIV/AIDS [24], accounting for approximately 40% of deaths globally [20]. HIV disease is equally a major global health concern causing substantial morbidity, mortality rate, human suffering, and development challenges [25]. The rapid disease progression and associated leading opportunistic infection, TB, contribute to the high mortality rates [26]. Beyond affecting the health of individuals, HIV and TB diseases are stigmatized [17] and have a significant impact on the social, economic and political stability of the hardest-hit countries [27,28].

Kenya suffers from the dual epidemics ranking 4^th^ and 15^th^ globally in the high disease burden for HIV and TB respectively [29–31]. The high HIV prevalence in Kenya is the major driver of TB related mortality and TB prevalence is also the leading cause of HIV related mortality [31]. Studies on TB and HIV incidences in Kenya have limited spatial and temporal scoping making generalizations of their findings to the whole country difficult. This study uses routine case notification data to provides more accurate estimates and insights on the elevated risk areas of HIV and TB individually and jointly over time. By using the spatiotemporal approach to modeling HIV and TB diseases simultaneously, we present the shared and disease-specific spatial risk patterns and explore their temporal evolution. The joint model also determines the combined and disease-specific elevated risk areas.

## Materials and methods

### Study location

The study was conducted in Kenya, a country of great diversity situated in East Africa extending between latitudes 4^0^30 □N and 4^0^30 □S and longitudes 34^0^00□E and 42^0^00□E [32]. Kenya has a coastline stretch of approximately 14420 km along the Indian ocean mostly covered by salt-tolerant mangrove trees that creates a distinctive ecological zone [33]. The total coverage area of Kenya is 583,367km^2^ with 569,140km^2^ as land area and 14,227km^2^ being water area [34]. The diversity of Kenya’s landscape is shaped by four distinguishable relief zones; these are the coastal and eastern broad plains, the central and western highlands, the Rift Valley and Lake Victoria basins [35].

Kenya is administratively subdivided into 47 counties as the first level of administrative subdivisions which in turn are further subdivided into 290 sub-counties and 1450 wards as the second and third levels of administrative subdivisions respectively [36]. The population of Kenya is unevenly distributed throughout the country and predominantly urban. The population density has remained on the increase from 77.9% in 2012 to 88.2% in 2017 per sq.km [37–39]. Supplementary information S2 presents the geospatial arrangements and the list of the 47 counties of Kenya according to their corresponding geographic codes as used in this study. The population estimates per county from 2012 – 2017 are in supplementary information S3.

A big obstacle to human capital development in Kenya is health challenges. Many people are exposed to a wide range of disease burdens largely because of the country’s geographical, economic, political and climatic conditions [40] which are further compounded by inadequate resources to mitigate the impact of health risks [41]. Presently, cases of emerging and re-emerging diseases – like TB and HIV - are on the rise thereby having important implications on public health policy processes [42]. Understanding variations in incidence trends at the county level – where health services are planned, organized and delivered – is essential in addressing health inequalities.

### Data Sources

This study considered routine case notification data on TB and HIV diseases in 47 counties of Kenya for 6 years, 2012-2017. Case notifications are data from specific subpopulations who seek treatment and care from health facilities; these are geographically representative of nearby populations. The data were collected and made available by two main sources; the National Tuberculosis Leprosy & Lung Disease Program (NTLD-P) and National AIDS & STIs Control Program (NASCOP). The Government of Kenya has routine case-based monitoring and reporting regulations for TB and HIV diseases through the NLTP and NASCOP programs. The Integrated Electronic Medical Records (EMR) Data Warehouse (IDWH) is the on-line case-based repository hosted by NASCOP accommodating all EMR databases from all health facilities across the country. It operates both as a repository and analytics platform presenting data through interactive dashboards and ad-hoc data analysis. Health facilities update their EMR databases into the IDWH on a monthly basis. The Tuberculosis Information from Basic Unit (TIBU) is the centrally located case-based surveillance system hosted by NLTP that allows for real-time reporting. Since its inception in 2012, TIBU has made notifications of TB patients very timely and instant in report generation. All public, faith-based and private treatment centers in the country enter data into the TIBU system. There are 301 TB control zones across the 47 counties of Kenya which are coordinated by the Sub County TB and Leprosy Coordinators (SCTLCs), who are responsible for notifying TB cases from health facilities in their control zones into the TIBU system. Both programs have adapted the data recording and reporting standards of WHO at the health facilities in every county and the national surveillance system. For the purpose of this study, we analyze the data aggregated per county per year for the period 2012-2017.

### Ethical considerations

This study involved the use of non-identifiable secondary data collected as part of the routine programs monitoring. Ethical permission to use the routinely collected data was obtained from NASCOP and NLTP, which are commissioned by the Ministry of Health to host the data surveillance systems for the HIV and TB programmes respectively. The study was subjected to Human Research Protection Review by the African Medical Research Foundation (AMREF Health Africa) Ethical and Scientific Review Committee (ESRC), which determined it not to constitute human participation.

### Hierarchical Model specification

We formulated a statistical model that combined the spatiotemporal methods to the modeling of HIV and TB simultaneously. The study applied a full Bayesian Hierarchical approach on the model to estimate the spatial and temporal parameters for the two diseases individually and jointly. We applied the spatial and temporal specifications of [9] and [43] to our model. While [9] mapped a single disease for two subpopulations, we mapped two diseases -HIV and TB-with co-epidemic overlap. They were also keen to interpret the unexpected differential risks between two subpopulations; our aim was to determine the shared and specific spatiotemporal patterns to interpret the relationships between the two diseases. Unlike [43], our model accounts for the joint space-time interaction using similar specifications as [9].

#### a. Log-linear model

The initial step in defining our model within the Hierarchical Bayesian framework was selecting the probability distribution for the observed data. This study utilized the Poisson distribution from the exponential family. For county s in the year t, we modeled cases notification y_dst_ for disease d, where d=1 is HIV and d=2 is TB as;

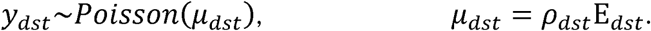

This study defined the mean μ_dst_ in terms of the unknown relative risk ρ_dst_ and the expected number of cases E_dst_. We computed E_dst_ per county per year. Our statistical consideration for the standard population N was the average of the pooled county population estimates, i.e. 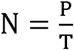, where P is the estimated population of county s, which we are considering to be the population at risk of disease d and T is the number of years, which is six years for this study. We calculated the crude rate as 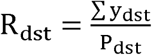, where ∑y_dst_ and P_dst_ are the number of cases for disease d and estimated population respectively for county s in the year t. We then multiplied the crude rate by the standard population to obtain the expected number of cases for disease d for county s in the year t as;

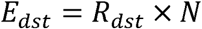

The linear predictor of the unknown relative risk was on the logarithmic scale, η_dst_ = log (ρ_dst_) which is the recommended invertible link function for the Poisson family of distributions. The variability of the cases around the unknown relative risks ρ_dst_ for HIV and TB respectively were as follows;

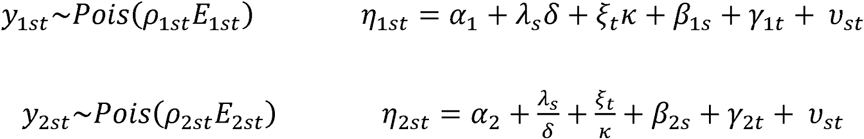

The linear predictor was defined by the following terms; the shared spatial effect, (**λ** ={λ_s_}_s=1,2,…,S_), the disease-specific spatial effect (**β** ={β_ds_}_d=1,2; s=1,2,…,S_), the shared time trend (**ξ** ={ξ_t_}_t=1,…,T_), the disease-specific time trend (**γ** ={γ_dt_}_d=1,2; t=1,…,T_), and the space-time interaction term (**ν** ={ν_st_}_s=1,…,S; t=1,…,T_). The notations α_d_, λ_s_, and ξ_t_ captured disease-specific intercept, space and time main effects respectively whereas γ_dt_ and β_ds_ were disease-time and disease-space interactions of order 2 respectively. The coefficients δ and κ represented the spatial and temporal scaling parameters on the shared term to the risk of TB compared to HIV. Even though the overall relative risk level is the same for both diseases, the magnitude of the area-specific and time-specific relative risks may differ - hence the need for the scaling parameters [9,44,45]. Therefore, contribution of the shared component to the overall relative risk is weighted by the scaling parameters to allow different risk gradients for each disease.

We applied a symmetric formulation to both the shared and disease-specific random effects, implying that λ_s_, and ξ_t_ captured the common spatial and temporal patterns. The terms γ_dt_ and β_ds_ allowed for departures from the shared patterns for the different diseases. The space-time interaction term, ν_st_ provided additional flexibility towards identifying varying patterns. We assumed the shared random effects to be the structured effects and the disease-specific random effects to be unstructured effects

#### b. Bayesian prior specification

In a Bayesian framework, random effects are unknown quantities assigned to prior distributions that reflect any prior knowledge on the structure of the effects. The model assigned priors and generated the posterior distribution used for deriving the conditional densities for posterior sampling.

The spatial random effects λ_s_ and β_s_ assumed a spatially correlated prior distribution (CAR spatial priors) with a neighborhood matrix W defined by contiguity; τ_λ_ and τ_β_ were the prior hyperparameters. The CAR spatial prior defines a binary specification for the geographical contiguity such that correlation is certain for geographically adjacent areas that is:

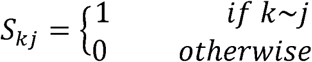

The non-contiguous areal units are conditionally independent given the values of the remaining random effects. To reflect the prior of yearly fluctuations for ξ_t_ and γ_dt_, the study assumed a random walk prior of order 1 (RW (1)) with a weighted matrix Q which defines the temporal neighborhood with τ_ξ_ and τ_γ_ as the prior hyperparameters. The space-time interaction term ν_st_ prior was a simple exchangeable hierarchical structure ν_st_ ∼ N (0, τ_ν_). We defined improper priors for the intercept as α ∼ N(0,0). The scaling parameters assume normal priors N(0, σ^2^) which are symmetric around zero, therefore, any value is as equally likely as the reciprocal value and the posterior distribution of the relative risks for each disease are exactly the same. For the distribution of the hyper-parameters, we assumed the default specifications of INLA whereby we assigned minimally informative priors on the log of the precision of both the structured and unstructured effects, logGamma(1, 0.0005). INLA estimates the posterior marginal distribution for the hyperparameters using an integration-free algorithm described in [46]. We use INLA approach for the model estimations as it is capable of handling complex models with large predictor spaces. Equally the approach does not require convergence checking (unlike McMC) [47,48] as it does not suffer from slow convergence and poor mixing.

## Results

To determine the areas of high risks, we created the spatial maps of the standardized incidence ratio for HIV (Fig 1) and TB (Fig 2). The two diseases displayed varying spatiotemporal patterns, though most of the regions of high risk for HIV were also high risk for TB. The progression of the risk during the period 2012 – 2017 was much faster in TB as compared to HIV

**Fig 1.**
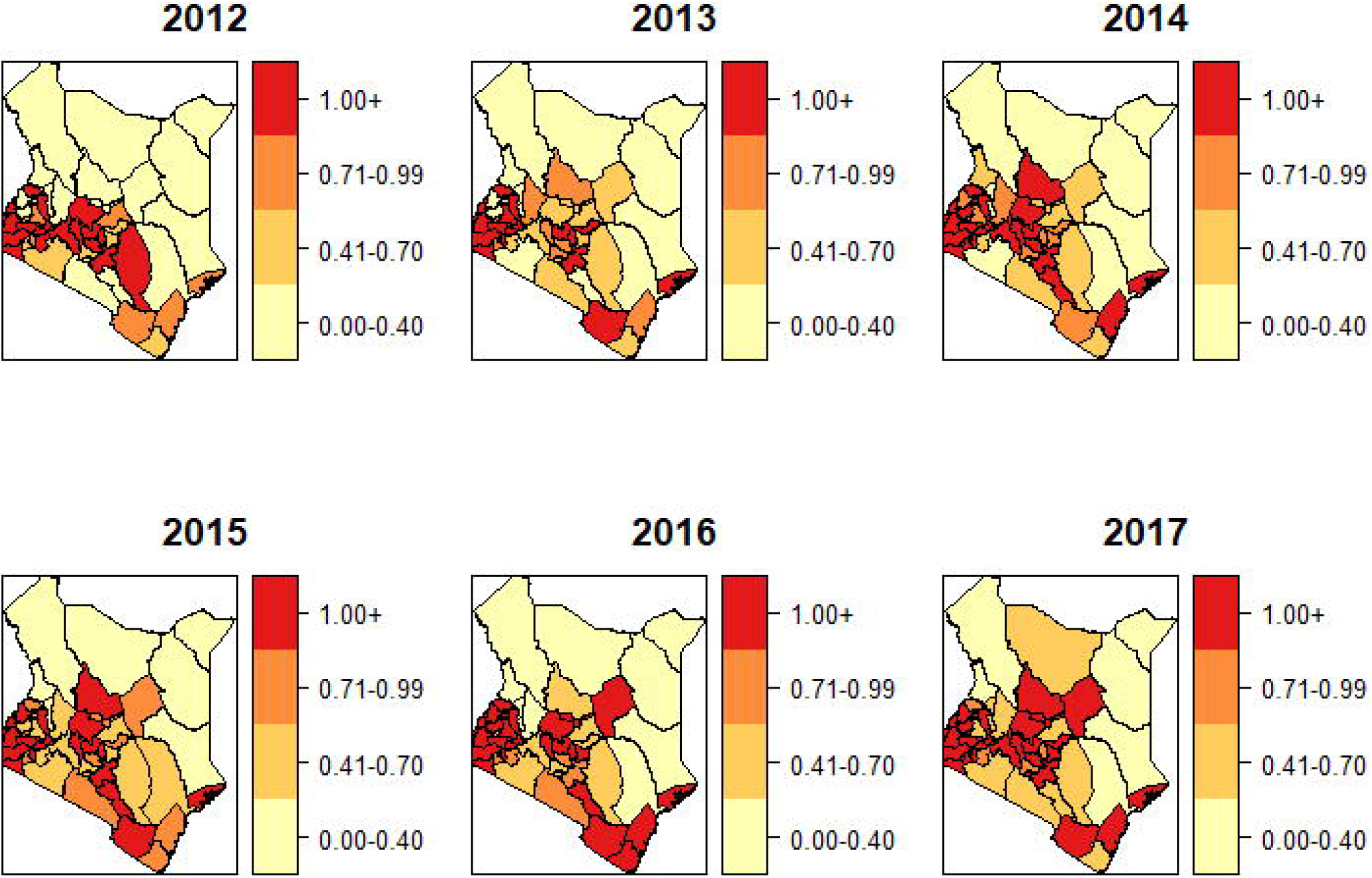
Standardized incidence ratio 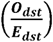 for HIV.

**Fig 2.**
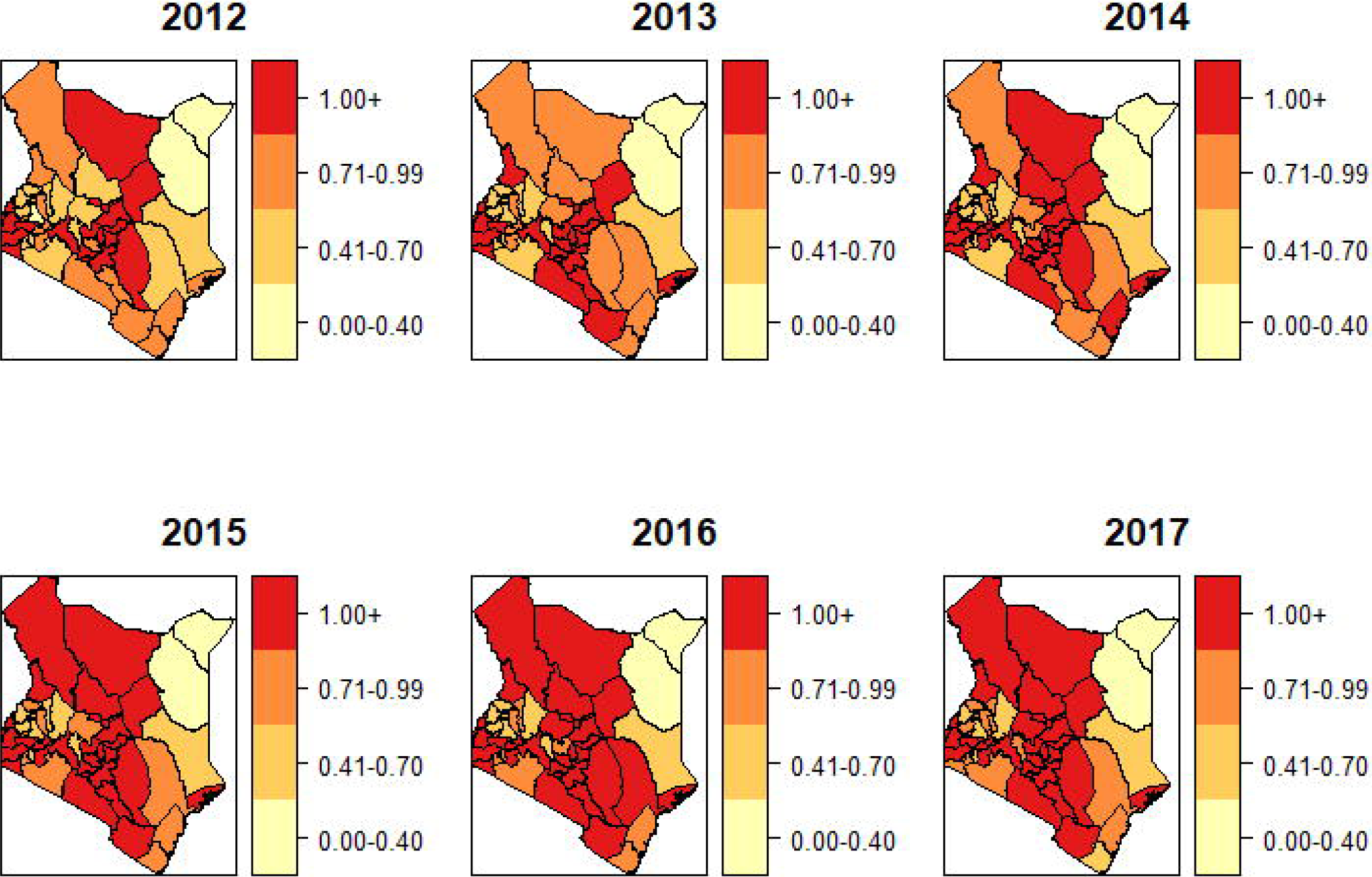
Standardized incidence ratio 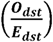 for TB.

We considered the analysis of the combined spatial patterns in the model. Table 1 provides the summary statistics of the shared and disease-specific spatial effects while Fig 3 presents the relative risk of the combined spatial patterns for HIV and TB. The estimates of the different spatial effects are statistically significant at 5% as depicted by the credible intervals. Both HIV and TB had similar areas of high risk in the South West, Central and South regions of Kenya. Further, TB revealed additional areas of high risk in the North West and North of Kenya.

**Table 1.**
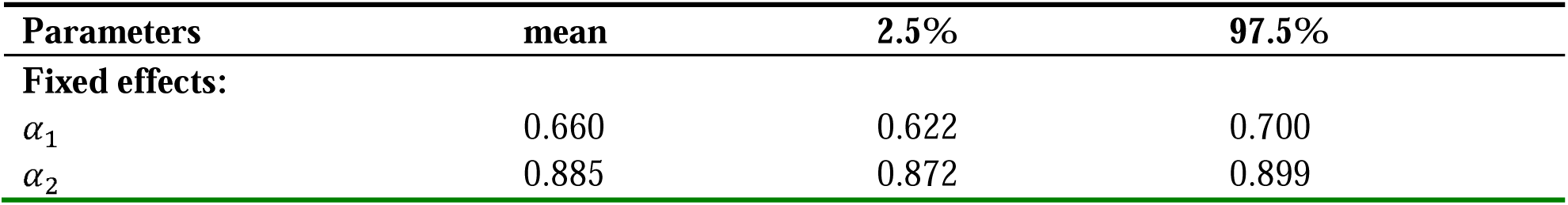
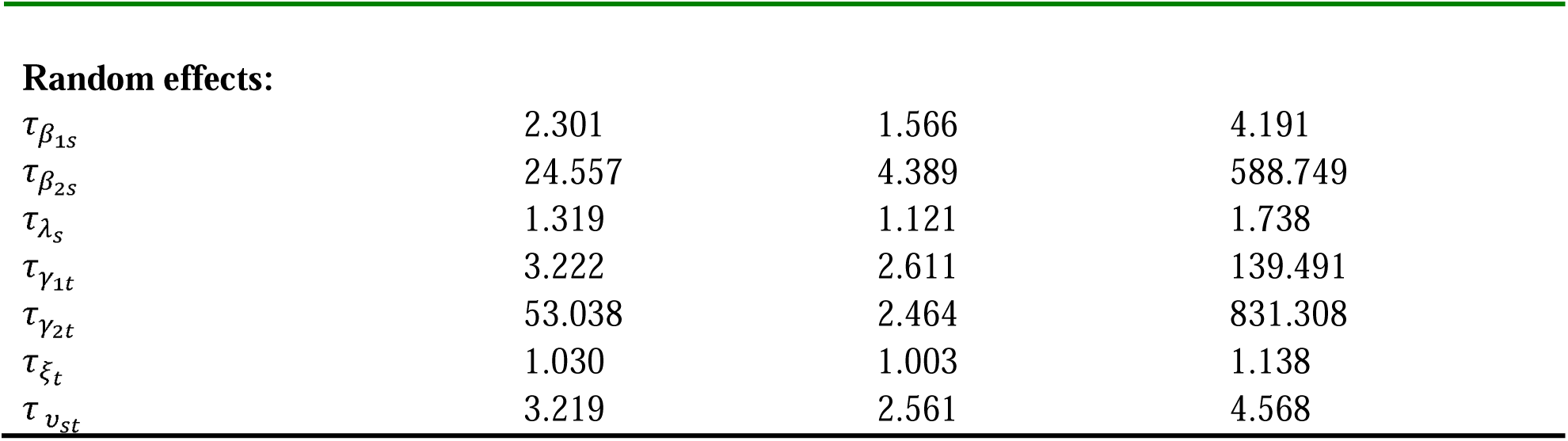
Summary statistics of the shared and disease-specific spatial and temporal effects

| Parameters | mean | 2.5% | 97.5% |
| --- | --- | --- | --- |
| **Fixed effects:** |  |  |  |
| $\alpha_1$ | 0.660 | 0.622 | 0.700 |
| $\alpha_2$ | 0.885 | 0.872 | 0.899 |
| **Random effects:** |  |  |  |
| $\tau_{\beta_{1s}}$ | 2.301 | 1.566 | 4.191 |
| $\tau_{\beta_{2s}}$ | 24.557 | 4.389 | 588.749 |
| $\tau_{\lambda_s}$ | 1.319 | 1.121 | 1.738 |
| $\tau_{\gamma_{1t}}$ | 3.222 | 2.611 | 139.491 |
| $\tau_{\gamma_{2t}}$ | 53.038 | 2.464 | 831.308 |
| $\tau_{\xi_t}$ | 1.030 | 1.003 | 1.138 |
| $\tau_{v_{st}}$ | 3.219 | 2.561 | 4.568 |

**Fig 3.**
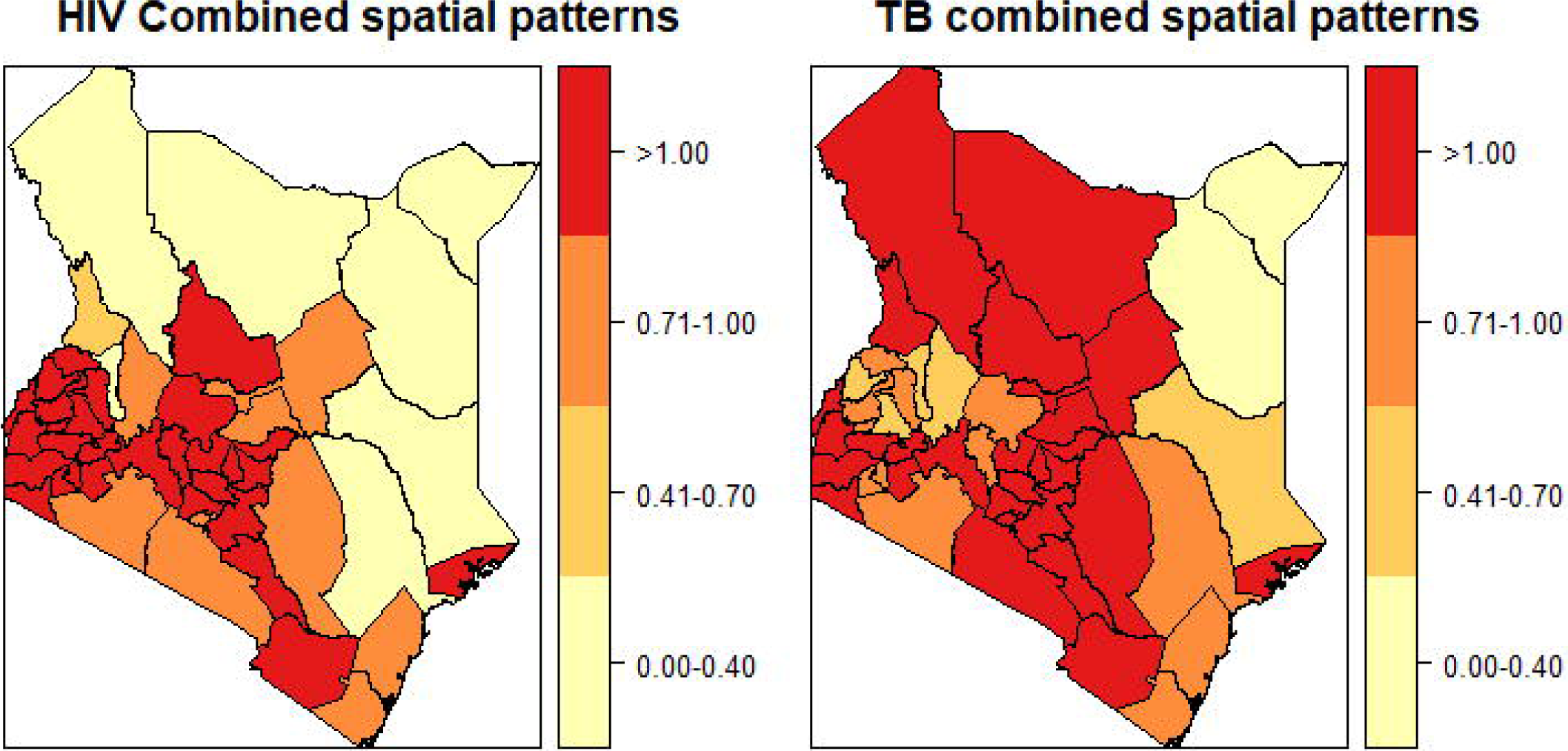
Relative risk of the combined spatial patterns (*λ*_*s*_ + *β*_*ds*_)

The posterior means of the shared and disease-specific spatial patterns are presented in Fig 4. In all the maps, common areas of high risk were in the west part of the country. Equally, the shared spatial pattern showed fewer regions of high risk compared to TB-specific spatial patterns. This could be because of higher dependence of HIV on the shared spatial term making the shared pattern to account for most of HIV spatial patterns

**Fig 4.**
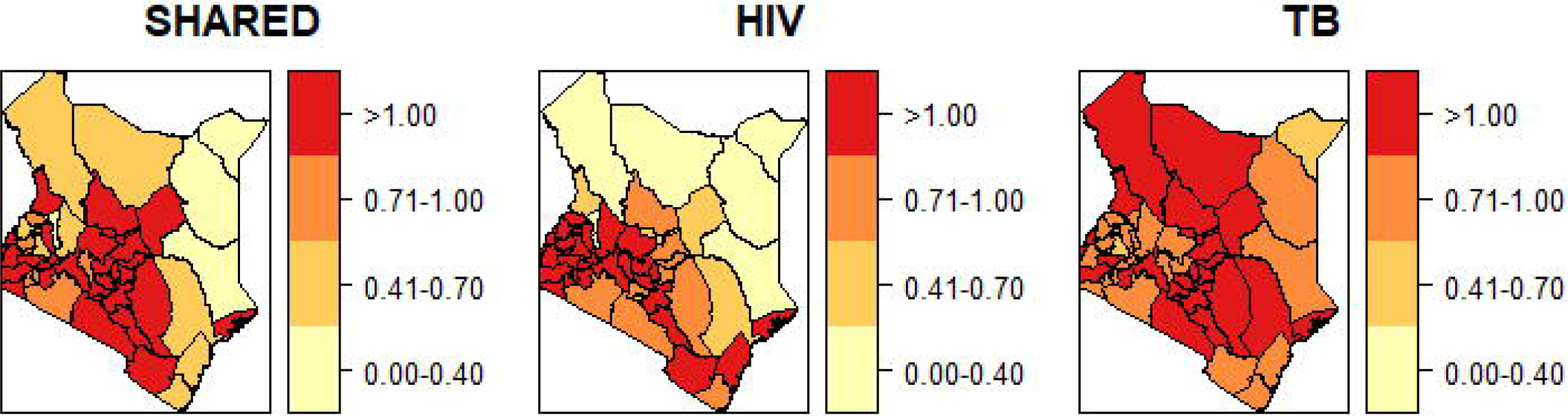
Relative risk of shared spatial patterns (*λ*_*s*_) and disease specific spatial patterns (*β*_*ds*_)

The posterior means of the shared, disease-specific, and combined temporal trends are in Fig 5. The shared temporal effect displayed the overall decreasing risk trend in time with estimates close to zero for most years. The HIV temporal trend exhibited increasing risk over time displaying relative risks between 0.8 - 1.2 whereas the TB temporal trend presented a nearly constant trend across the years with relative risks close to one. The combined temporal trends for HIV and TB showed an increasing risk trend for both diseases. From the combined temporal trends graph, HIV risk was relatively lower than TB in 2012 and 2013. From 2015 - 2017, the HIV risk trend surpassed that of TB. Even though the shared effect shows minimal evidence on the joint diseases, the disease-specific and combined temporal effects show very similar temporal pattern.

**Fig 5.**
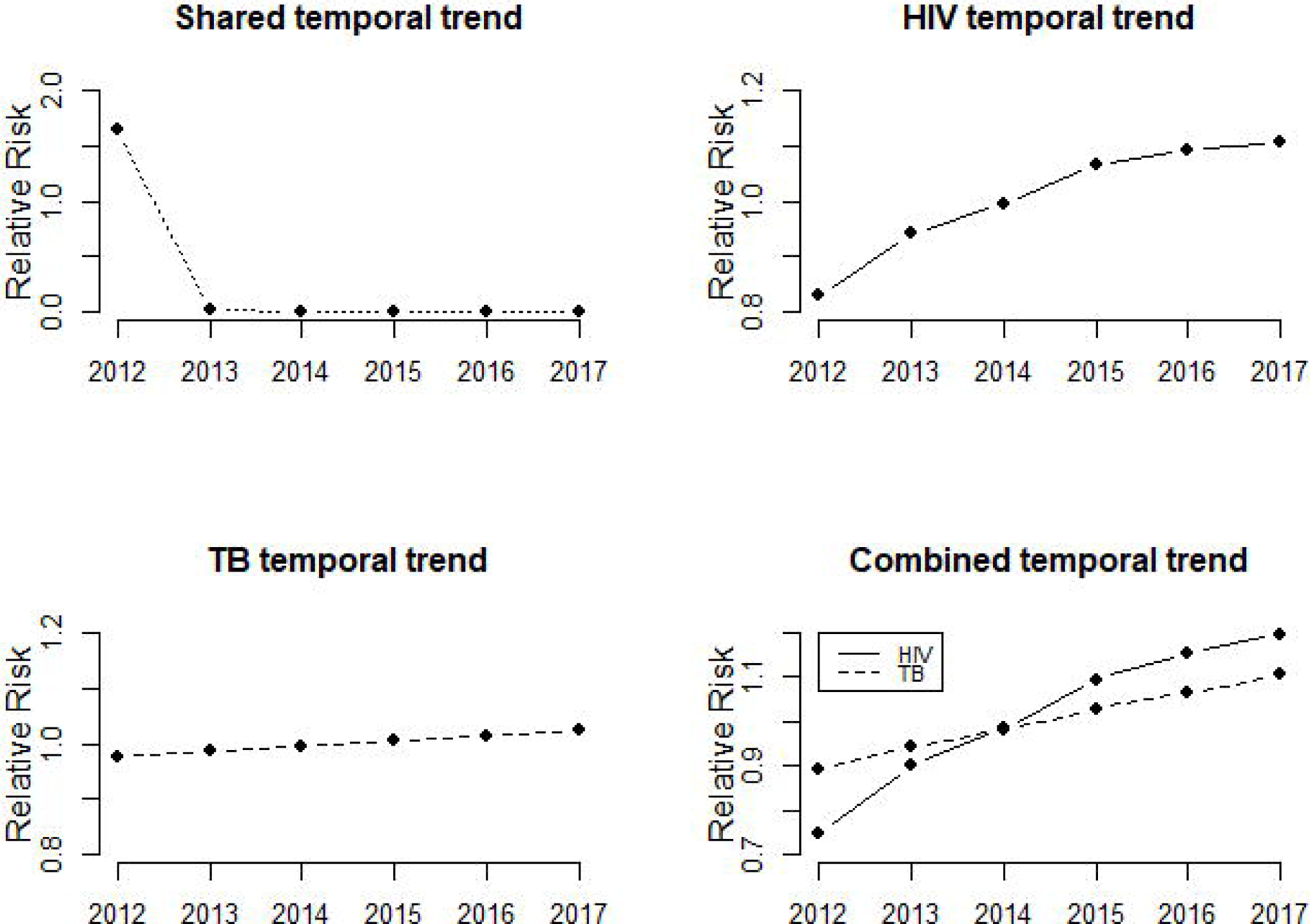
Relative risk of the shared *ξ*_*t*_, specific *γ*_*dt*_ and the combined temporal trends (*ξ*_*t*_ + *γ*_*dt*_)

The posterior probabilities for the smoothed joint Spatio-temporal interaction 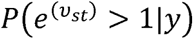 are in Fig 6, and the posterior estimates are presented in Table 1. Over the years the joint incidence risk reduced progressively with all spatial regions having a relative risk less than 1.

**Fig 6.**
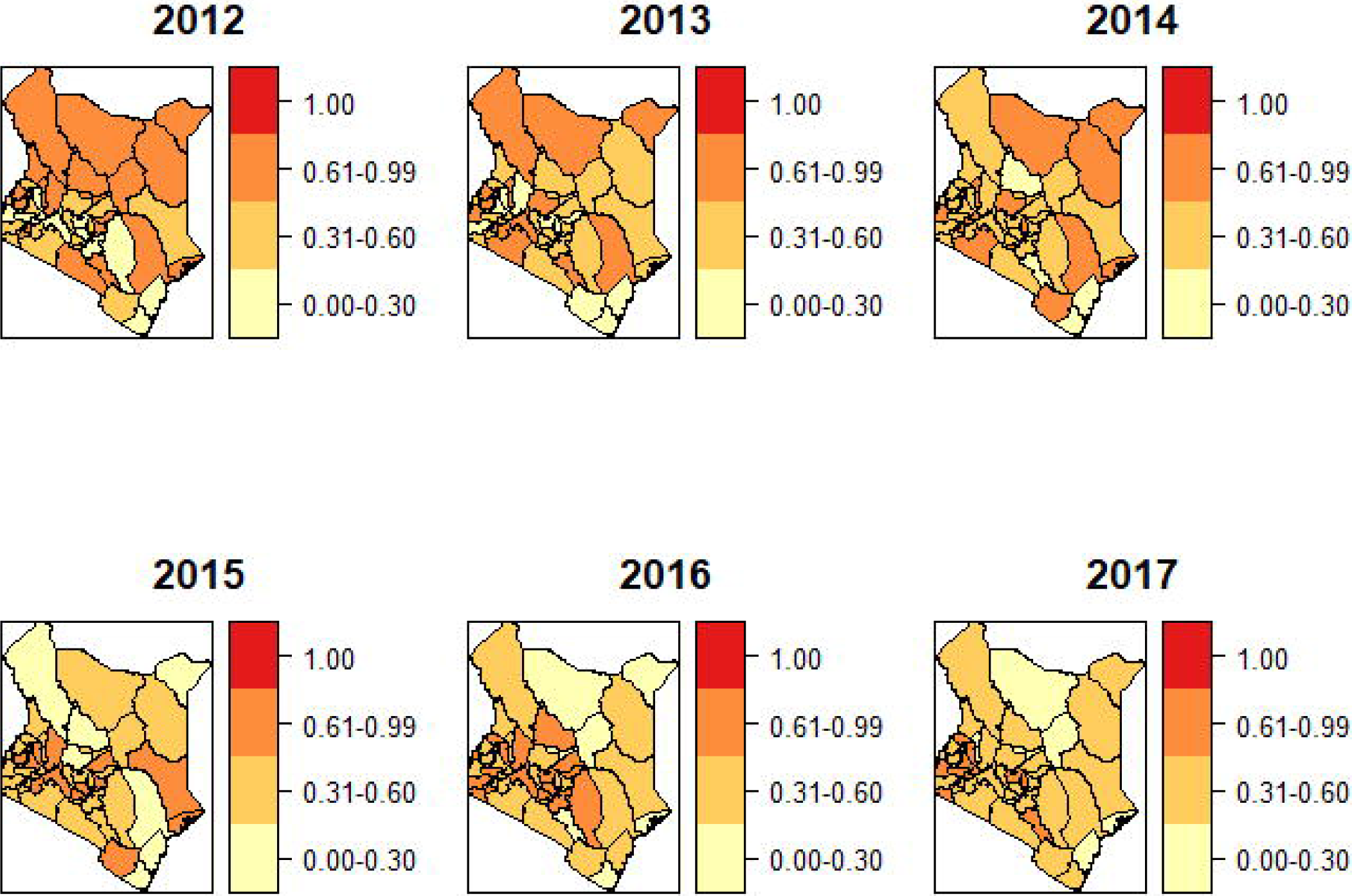
Posterior probabilities for the joint Spatio-temporal interaction 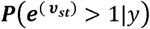.

Fig 7(a) displays the spatial pattern of the posterior mean for the joint disease risk 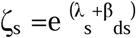. The uncertainty associated with the posterior means, which lays in the exceedance risk ζ_s_: P (ζ_s_>1|y), is presented in Fig 7(b). The extreme West and Central regions depicted elevated joint relative risks characterized by a spatial relative risk above one and exceedance probability above 0.8

**Fig 7.**
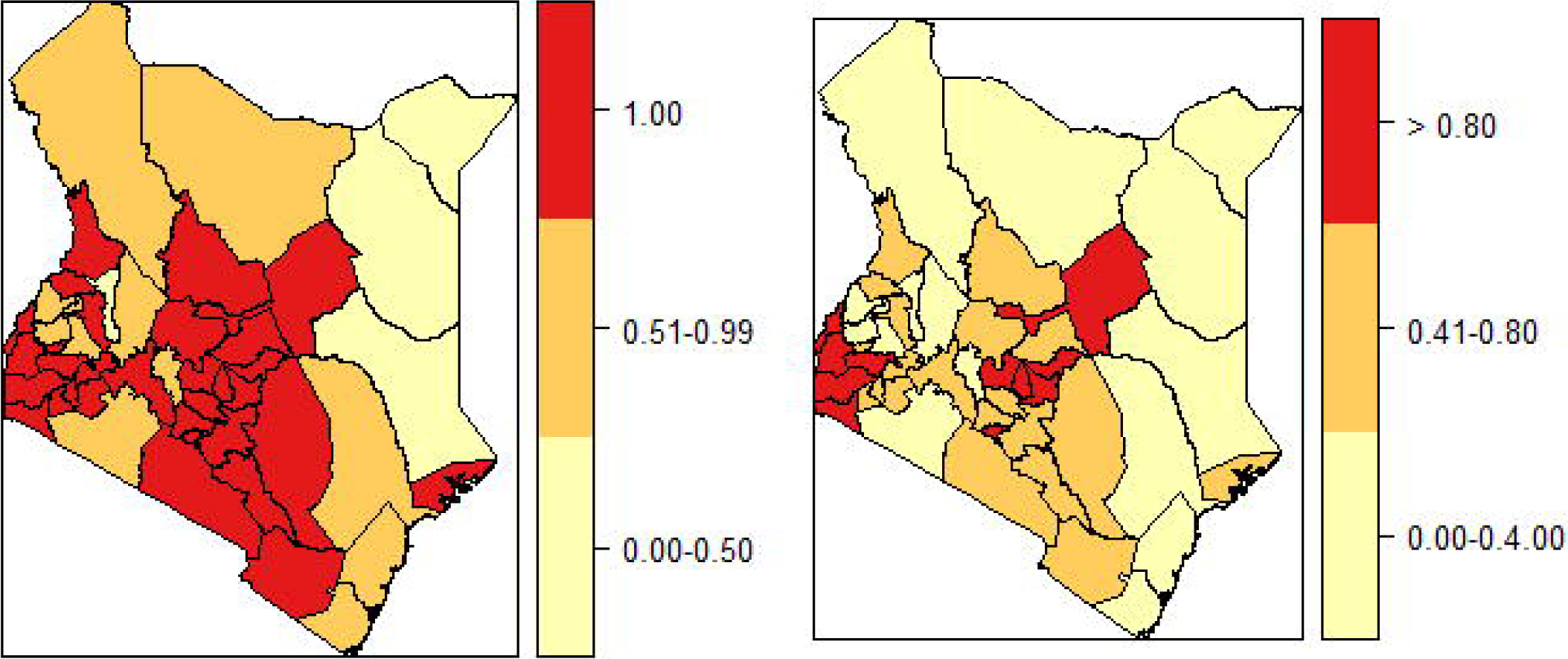
Maps of the spatial pattern of joint disease risk 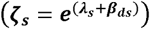 and uncertainty for the spatial effect (*ζ*_*s*_: *P* (*ζ*_*s*_>1|*y*))

## Discussion

We presented a Bayesian Hierarchical approach to the joint modeling of spatio-temporal routinely collected health data. The joint modeling approaches have yielded substantial co-dynamic insights via mathematical, statistical and computational approaches [49]. By optimizing the spatial scale at different points in time, spatial heterogeneity influences the interpretation of temporal patterns more especially in disease dynamics and surveillance [49]. This is especially true for the case of HIV and TB that have significant geographic overlap and are subject to diverse regional variations in their co-dynamics. HIV and TB rank as the leading causes of death from infectious diseases globally with an estimated 2.5 million new HIV infections and 8.7 million incidences of TB annually [50,51]. They have a close link even though their biological co-existence and co-dynamics vary regionally with much burden in Sub-Saharan Africa [31,52]. This study determined the space-time joint risk trends of HIV and TB in Kenya. Our model enabled us to define the shared and specific spatial and temporal patterns of HIV and TB thus identifying similarities and differences in the distribution of the relative risks associated with each disease. The model separately estimated the shared and disease-specific relative risks and displayed the spatial-disease, temporal-disease, and spatio-temporal disease interaction effects across all regions. We included scaling components on the shared spatial and temporal parameters to compare their strength signals for HIV and TB.

The disease-specific spatial and temporal patterns detected areas with varying spatial trends and temporal variations for each disease. The HIV high-risk areas were to the further west of Kenya spreading towards the central and further south. The TB high-risk areas were similar to the HIV high-risk areas but also spread upwards towards the North. The TB geographical progression in relation to HIV was proportionally higher which could reflect environmental factors favoring the TB spread in the high-density settlements especially in the towards the North. These findings are corroborated in other studies by [31,53]. Looking beyond Kenya, studies by [54] revealed that TB appeared to outpace HIV in Rwanda and Burundi while HIV greatly outpaced TB in Mauritania, Senegal and The Gambia

Joint temporal analysis is important when investigating the temporal coherency of epidemiological trends from the same area [54]. In our study, the shared temporal trend presented a steep decrease from 2012 to 2013 then an almost constant risk without any significant variation over time. However, the disease-specific and combined temporal trends presented an increased risk over time. The temporal trend of HIV risk was lower than that of TB for the years 2012 and 2013 but between 2015 and 2017 the HIV risk was higher than TB risk. Similar studies in Sub-Saharan Africa that utilized routinely collected data observed similar temporal dynamics [55–58]. A possible explanation could be HIV drives TB related incidences, therefore, the incidence and prevalence of TB increases (decreases) with increasing (decreasing) HIV trends [59–61].

Our study successfully detected the spatial congruence in the distribution of TB and HIV in approximately 29 counties around the western, central and southern regions of Kenya. The spatial patterns were largely similar for Homabay, Siaya, Kisumu, Busia and Migori counties as the high risk with Mandera, Wajir and Garissa counties at low risk for both HIV and TB. The distribution of the shared relative risks had minimal difference with the HIV disease-specific relative risk whereas that of TB presented many more counties as high-risk areas. This could be attributed to higher dependence of HIV on the shared spatial term making the shared pattern account for most HIV spatial patterns. Similar studies by [62] in China and [53] in Uganda observed significantly persistent clusters for TB and HIV using the spatial co-clustering approach. They examined the clusters exhibited by each disease as well as the combined.

Our model also presented areas of elevated joint risk by looking at the posterior probabilities of the relative risk to detect the joint geographical patterns. There was strong evidence of disease-time, disease-space and space-time interactions. Studies by [62] and [63] also found a strong association on the joint risks of HIV and TB they used bivariate maps to show that the joint distribution for both TB and HIV diseases was spatially heterogeneous across Brazil.

The primary limitation of the study is using routine case notification data as a surrogate measure of the general population of infected persons; this type of data has challenges of underreporting. The spatial regions were based on the counties since there was no data for sub-county or health facility levels. The age and sex standardized rates were not utilized as the HIV data was not disaggregated by age and gender. Whereas this kind of data is not completely spatially random for the joint epidemic burden, it still captures the spatiotemporal patterns of incidence risk, which is the ultimate goal of this study.

## Conclusions

We defined a joint Bayesian space-time model of two related diseases to jointly quantify the risk of TB relative to HIV thereby facilitating the comparative benefits obtained across populations. The disease burden was apparent at each spatial level of analysis. Identifying the spatial and temporal similarities between HIV and TB enabled us to understand the shared risk. The flexibility and informative outputs of Bayesian Hierarchical Models played a key role in clustering these risk areas. Quantifying how HIV and TB varied together provided additional insights towards collaborative monitoring of the diseases and control efforts. To control the HIV-TB twin epidemic it is important to determine the TB life history that could greatly reduce TB incidences if intervened and which stages of HIV needs optimized TB control benefits

## Data Availability

The HIV and TB data used in this study are available from NASCOP (P.O. Box 19361-00202, Nairobi-Kenya; telephone: +254-775597297; https://dwh.nascop.org/) and NLTP (P.O. Box 20781-00202, Nairobi-Kenya; telephone: +254-773977440; http://pms.dltld.or.ke/). Interested researchers may contact Dr. Catherine Ngugi for the HIV data and Dr. Elizabeth Onyango for the TB data. The county population estimates for the period 2012-2017 are available from the Keya National Bureau of Statistics (P.O. Box 30266-00100, Nairobi-Kenya; telephone: +254-20-317583; www.knbs.or.ke).

## Acknowledgments

The authors sincerely acknowledge the National Tuberculosis, Leprosy and Lung Cancer Program (NLTP) and the National AIDS and STI Control Program (NASCOP) for their cordial support in providing the data used in this study

## Supporting Information

S1 Text. R codes used in the analysis

S2 Text. List of counties in Kenya

S3 Text. County population estimates 2012-2017

## Notes

### Competing Interest Statement

The authors have declared no competing interest.

### Funding Statement

The authors would like to thank the African Union and the Pan African University Institute of Basic Sciences Technology and Innovation (PAUISTI) for funding the study through VO’s Ph.D. work

